# Cardiac Safety and Hemodynamic Effects of Mannitol Compared with Hypertonic Saline in Intracranial Hypertension: A Systematic Review and Meta-analysis

**DOI:** 10.1101/2025.11.06.25339663

**Authors:** Farzan Fahim, Farbod Tabasi Kakhki, Seyyed Mohammad Hosseini Marvast, Sana Mahdian Rizi, Sadra Abedinzadeh, Reihane Qahremani, Ali salmani, Mohadese Ahmadloo, Mobina ghamarpour, Mohammadsadegh Jafari, Sayeh Oveisi, Alireza Zali, Saeed Oraee Yazdani

**Affiliations:** Neurosurgery resident at Shohada-E-Tajrish Hospital, Shahid Beheshti University of Medical Science, Tehran Iran; Functional Neurosurgery Research Center, Research Institute of Functional Neurosurgery, Shohada Tajrish Comprehensive Neurosurgical Center of Excellence, Shahid Beheshti University of Medical Sciences, Tehran, Iran; professor of Neurosurgery, Shohada-E-Tajrish Hospital, Shahid Beheshti University of Medical Sciences, Tehran, Iran; Associated professor of neurosurgery in Shohada-E-Tajrish Hospital, Shahid Beheshti University of Medical Sciences, Tehran, Iran

**Keywords:** mannitol, intracranial pressure, hypertonic saline, cardiac complications, arrhythmia, hemodynamics, electrolytes, meta-analysis

## Abstract

**Background:** Mannitol is widely used for reducing intracranial pressure (ICP), yet concerns persist regarding its cardiovascular safety. This systematic review and meta-analysis evaluated cardiac adverse events and mortality associated with mannitol compared with hypertonic saline (HS) in adults with intracranial hypertension.

**Methods:** Following PRISMA 2020, nine studies (five RCTs, two cohorts, one open-label trial, and one case-control study; n = 428) comparing mannitol with HS or standard ICP therapy were analyzed. Random-effects models generated pooled risk ratios for cardiac events and mortality. Risk of bias was assessed using JBI tools.

**Results:** Mannitol consistently reduced ICP by 20–35% within 30 minutes and caused only transient decreases in MAP (5–8 mmHg). Cardiac adverse events did not increase significantly (RR 1.77; 95% CI 0.00–6.3×10⁵; p = 0.67; I² = 42.5%). Mortality analysis (four studies, n = 150) showed a non-significant numerical elevation (RR 3.6; 95% CI 0.83–15.6; p = 0.06), driven mainly by one small outlier trial. Electrolyte shifts and QTc changes were more closely linked to hyperosmolality than to the specific agent used.

**Conclusion:** Mannitol is effective for ICP reduction without significant increases in cardiac or mortality risk compared with HS. Cardiac vulnerability appears exposure-dependent, reinforcing the need for conservative serum osmolality targets (<320 mOsm/kg), osmolar-gap monitoring, and structured ECG and electrolyte surveillance. Larger multicenter trials with standardized cardiac endpoints are required.

## Introduction

Intracranial pressure (ICP) reflects the equilibrium among brain parenchyma, cerebral blood volume, and cerebrospinal fluid (CSF) within the cranial vault (1, 2). In adults, normal ICP ranges from 5 to 15 mmHg (3), while intracranial hypertension (IH) is defined as sustained pressures exceeding 20 mmHg (4, 5). IH arises in different neurological conditions, including cerebral edema (6), traumatic brain injury (TBI), infections, intracranial hemorrhage, hydrocephalus, ischemia–reperfusion injury, and hypertensive crises (7–12), which are strongly associated with brain herniation, cerebral hypoxia, coma, and mortality (2, 5, 8, 10, 13). Rapid and targeted ICP reduction is important to minimize secondary brain injury (13). Age-related vascular stiffening, impaired CSF circulation, and reduced compensatory mechanisms also can cause severe IH in the elderly (14–16), which points to safe and effective hyperosmolar therapies.

Hyperosmolar therapy, primarily mannitol and hypertonic saline (HS), remains a cornerstone of ICP management (10, 17). Mannitol, a 6-carbon inert sugar alcohol approved by the FDA, reduces ICP by increasing plasma osmolality and promoting water efflux (5, 10, 18, 19). It is routinely used to treat cerebral edema following brain insults and neurosurgical procedures (20, 21). Despite its widespread use, some concerns persist due to its systemic effects. Case reports and small clinical series have described complications such as transient heart failure triggered by rapid intravascular volume shifts (18), impaired cardiac function in the elderly (22), hypotension, dehydration, acute kidney injury, and hypernatremia (5, 20, 23). Mannitol-induced diuresis may also cause hypovolemia in vulnerable patients (24). However, these observations derive from heterogeneous and old studies that were done before the use of modern hemodynamic monitoring.

Comparative research over the past decade has increasingly evaluated mannitol alongside HS across neurosurgical, TBI, stroke, and acute liver failure populations (25–32). These studies consistently demonstrate effective ICP reduction, typically 20–35% within the first 30 minutes of infusion, though with variable impacts on mean arterial pressure (MAP), cerebral perfusion pressure (CPP), and electrolytes (chunk 2; 25–32). The hemodynamic effects of mannitol versus HS remain clinically relevant, given that transient MAP decreases of approximately 5–8 mmHg have been documented, and both hyperosmolar agents influence plasma osmolality with potential downstream cardiac effects (27, 28, 32). Notably, QTc prolongation and atrial arrhythmias appear more closely associated with hyperosmolality itself rather than with the specific osmotic agent used (32).

Despite these considerations, direct cardiac outcome reporting has been inconsistent. Among the nine studies synthesized in the present review, only three, Bilotta et al. 2012, Su et al. 2021, and Hasanpour Mir et al. 2012, systematically compared cardiovascular or pulmonary complications between mannitol and HS. Existing data suggest potential exposure-dependent vulnerability in cardiac risks, but these findings remain insufficient to determine whether mannitol has distinctive cardiac risks relative to HS. Furthermore, prior meta-analyses have prioritized ICP control and brain relaxation (33–37) or TBI-specific endpoints (35, 36, 38–40), leaving uncertainty regarding differential cardiac and hemodynamic safety profiles across diverse IH etiologies.

Based on the necessity of cardiovascular stability in neurocritical care and the limited, uncertain reporting of cardiac outcomes in existing trials, a comprehensive synthesis is needed. Therefore, this systematic review and meta-analysis aim to evaluate the cardiac safety and hemodynamic effects of mannitol compared with HS in adult patients with intracranial hypertension, integrating evidence across randomized and observational studies to contextualize arrhythmias, QTc changes, electrolyte disturbances, MAP/CPP responses, ICP reduction, and mortality.

## Methodology

### Study Design

This systematic review was conducted in accordance with the Preferred Reporting Items for Systematic Reviews and Meta-Analyses (PRISMA) 2020 guidelines (Page et al., 2021). The protocol was prospectively registered in the International Prospective Register of Systematic Reviews (PROSPERO) to ensure methodological transparency and prevent duplication of efforts. The registration is publicly available in PROSPERO (Registration ID: CRD420251160241, registered on 02 October 2025).

A methodological revision was made on 22 October 2025, broadening the age inclusion criteria due to an insufficient number of studies meeting the original restriction. This modification aimed to enhance the comprehensiveness of the review while maintaining alignment with the original objectives.

### Search Strategy

A comprehensive literature search was conducted in October 2025 across the following electronic databases: PubMed, Scopus, Web of Science, Embase, and the Cochrane Library. No restrictions were applied regarding language or publication date to ensure broad coverage of relevant literature. Articles published in languages other than English were to be translated if included, although no such articles were ultimately retrieved.

The search included the following keywords: “mannitol,” “intracranial pressure,” “ICP elevation,” “cardiac effect,” “traumatic brain injury,” and “brain tumor.” Boolean operators (AND, OR) were used to refine and combine the terms.

An example of the PubMed search query is presented below:

(“Mannitol”[Mesh] OR mannitol[tiab] OR “Osmofundin”[tiab] OR “Osmitrol”[tiab] OR “(L)-Mannitol”[tiab]) AND (“Intracranial Pressure”[Mesh] OR “Hypertension, Intracranial”[tiab] OR “Elevated ICP”[tiab] OR “ICP, Elevated”[tiab] OR “Intracranial Pressure, Elevated”[tiab] OR “ICP Elevation”[tiab] OR “ICP Increase”[tiab] OR “Intracranial Pressure Increase”[tiab] OR “Brain Edema”[Mesh] OR “Brain Edema”[tiab] OR “Brain Injuries, Traumatic”[Mesh] OR “Brain Injury, Traumatic”[tiab] OR “Traumatic Brain Injury”[tiab] OR “Traumatic Brain Injuries”[tiab] OR “Brain Trauma”[tiab] OR “Trauma, Brain”[tiab] OR “Encephalopathy, Traumatic”[tiab] OR “TBI”[tiab] OR “TBIs”[tiab] OR “Brain Neoplasms”[Mesh] OR “Brain Neoplasm”[tiab] OR “Neoplasm, Brain”[tiab] OR “Brain Tumors”[tiab] OR “Brain Tumor”[tiab] OR “Tumor, Brain”[tiab] OR “Neoplasms, Brain”[tiab] OR “Brain Cancer”[tiab] OR “Brain Cancers”[tiab] OR “Cancer, Brain”[tiab] OR “Neoplasms, Brain, Malignant”[tiab] OR “Brain Neoplasms, Malignant”[tiab] OR “Brain Neoplasm, Malignant”[tiab] OR “Malignant Brain Neoplasm”[tiab] OR “Malignant Brain Neoplasms”[tiab] OR “Cancer of Brain”[tiab] OR “Cancer of the Brain”[tiab] OR “Malignant Neoplasms, Brain”[tiab] OR “Brain Malignant Neoplasm”[tiab] OR “Brain Malignant Neoplasms”[tiab] OR “Malignant Neoplasm, Brain”[tiab] OR “Malignant Primary Brain Tumors”[tiab] OR “Brain Neoplasms, Malignant, Primary”[tiab] OR “Brain Neoplasms, Primary Malignant”[tiab] OR “Malignant Primary Brain Neoplasms”[tiab] OR “Primary Malignant Brain Neoplasms”[tiab] OR “Primary Malignant Brain Tumors”[tiab] OR “Neoplasms, Intracranial”[tiab] OR “Intracranial Neoplasm”[tiab] OR “Intracranial Neoplasms”[tiab] OR “Benign Neoplasms, Brain”[tiab] OR “Benign Neoplasm, Brain”[tiab] OR “Brain Benign Neoplasm”[tiab] OR “Brain Benign Neoplasms”[tiab] OR “Brain Neoplasms, Benign”[tiab] OR “Benign Brain Neoplasm”[tiab] OR “Benign Brain Neoplasms”[tiab] OR “Brain Neoplasm, Benign”[tiab] OR “Neoplasms, Brain, Benign”[tiab] OR “Brain Tumor, Primary”[tiab] OR “Primary Brain Tumor”[tiab] OR “Primary Brain Tumors”[tiab] OR “Primary Brain Neoplasms”[tiab] OR “Brain Neoplasms, Primary”[tiab] OR “Primary Brain Neoplasm”[tiab] OR “Brain Neoplasm, Primary”[tiab] OR “Neoplasms, Brain, Primary”[tiab] OR “Brain Tumor, Recurrent”[tiab] OR “Brain Tumors, Recurrent”[tiab] OR “Recurrent Brain Tumor”[tiab] OR “Recurrent Brain Tumors”[tiab] OR “Brain Metastases”[tiab] OR “Brain Metastase”[tiab]) AND (“Arrhythmias, Cardiac”[Mesh] OR “Arrhythmia, Cardiac”[tiab] OR arrhythmia*[tiab] OR “Cardiac Dysrhythmia”[tiab] OR dysrhythmia*[tiab] OR “Atrial Fibrillation”[Mesh] OR “Atrial Fibrillation”[tiab] OR “Auricular Fibrillation”[tiab] OR “Paroxysmal Atrial Fibrillation”[tiab] OR “Familial Atrial Fibrillation”[tiab] OR “Ventricular Tachycardia”[Mesh] OR “Ventricular Tachycardia”[tiab] OR “Ventricular Tachyarrhythmia”[tiab] OR “Nonsustained Ventricular Tachycardia”[tiab] OR “Idiopathic Ventricular Tachycardia”[tiab] OR “Paroxysmal Supraventricular Tachycardia”[tiab] OR “Heart Failure”[Mesh] OR “Heart Failure”[tiab] OR “Congestive Heart Failure”[tiab] OR “Right-Sided Heart Failure”[tiab] OR “Left-Sided Heart Failure”[tiab] OR “Pulmonary Edema”[Mesh] OR “Pulmonary Edema”[tiab] OR “Wet Lung”[tiab] OR “Wet Lungs”[tiab] OR “Myocardial Infarction”[Mesh] OR “Myocardial Infarction”[tiab] OR “Heart Attack”[tiab] OR “Myocardial Infarct*”[tiab] OR “Ischemic Heart Disease”[tiab] OR “Myocardial Ischemia”[tiab] OR “Ischemic Heart Diseases”[tiab] OR “Cardiac Arrest”[Mesh] OR “Cardiac Arrest”[tiab] OR “Heart Arrest”[tiab] OR “Asystole”[tiab] OR “Cardiopulmonary Arrest”[tiab] OR “Hypotension”[Mesh] OR “Hypotension”[tiab] OR “Low Blood Pressure”[tiab] OR “Vascular Hypotension”[tiab] OR QTc[tiab] OR “QT interval”[tiab] OR troponin[tiab]).

The complete search strategies for all databases are provided in Supplementary File1.

Additionally, the reference lists of all eligible studies and relevant review articles were manually screened, leading to the identification of one additional article that was not captured in the initial search but met the inclusion criteria and was included in the final analysis.

### Eligibility Criteria

#### Population

Human participants with elevated intracranial pressure (ICP), irrespective of the underlying etiology (e.g., traumatic brain injury, stroke, intracranial hemorrhage, or brain tumor).

#### Intervention/Comparator

Mannitol administration to reduce ICP.

#### Outcomes

Quantitative measures of ICP reduction, mortality, and adverse effects, with an emphasis on cardiac outcomes: arrhythmias, ECG abnormalities, cardiac enzyme changes, electrolyte disturbances, and cardiovascular mortality.

#### Study Design

Randomized controlled trials (RCTs), cohort studies, or case-control studies.

### Exclusion Criteria

Studies were excluded if they met any of the following conditions:

1. Case reports or case series;
2. Reviews, meta-analyses, conference abstracts, or book chapters;
3. Non-human or in vitro studies;
4. Studies without available full text which cannot be found.
5. Studies not reporting relevant outcomes, particularly cardiac outcomes related to mannitol or hypertonic saline use;
6. Studies not meeting the age criteria, though the age limit was broadened during protocol revision (see PROSPERO registration CRD420251160241, revised 22 October 2025) to include all adult participants due to limited available evidence.

### Screening and Selection

A total of 2,121 records were initially identified through database searches (PubMed: 85; Scopus: 783; Web of Science: 65; Cochrane Library: 29; Embase: 1,159). All records were imported into Rayyan for systematic screening and duplicate removal, yielding 1,267 unique publications for assessment.

Title and abstract screening were independently performed by SA, AS, and SM. Any discrepancies were resolved through discussion, and unresolved conflicts were adjudicated by a fourth reviewer (FF). To systematically document exclusions during this phase, an Excel-based exclusion sheet (developed by FF) was used to record excluded studies, including article title, first author, country of origin, year of publication, DOI, and reason for exclusion (see Supplementary File 2,3).

After title and abstract screening, 54 studies were retained for full-text screening and recorded in the inclusion sheet. Additionally, manual screening of reference lists identified one eligible study, which was subsequently included in the final analysis.

Full-text screeniFull-text screening for eligibility was independently performed by SM and MH. Disagreements were resolved through discussion or consultation with a third reviewer (FT). Ultimately, nine studies met all inclusion criteria. These studies were selected for data extraction.

### Data Extraction

A standardized data extraction form was developed by FF. Data extraction was independently performed by MH and AS. The following variables were extracted:

- Study characteristics: first author, publication year, DOI, funding source, study region, study design, center type (single or multicenter), study period, ethical approval number, conflict of interest statement, blinding method, and risk of bias.
- Participant characteristics: sample size, age, sex distribution, baseline Glasgow Coma Scale (GCS), diagnosis, comorbidities, baseline ICP, past medical history, history of cranial surgery, baseline cardiac status, serum electrolytes, hemodynamic state, cardiac function monitoring, ECG findings, use of concomitant diuretics or vasoactive drugs, and potential confounding factors.
- Intervention details: type, concentration, dosage, and route of administration of mannitol or hypertonic saline, as well as duration and frequency of administration.
- Outcomes: ICP reduction, neurological improvement, adverse effects (cardiac and non-cardiac), including type, time of onset, incidence, duration, severity, and consequences, and mortality. Particular attention was given to cardiac outcomes such as arrhythmias, ECG changes, cardiac enzyme alterations, electrolyte disturbances, and cardiovascular-related mortality.

A detailed summary of all extracted variables (e.g., pre- and post-intervention ICP, GCS, cardiac parameters, electrolyte changes, and adverse event characteristics) is provided in Supplementary File 4.

### Risk of Bias

The methodological quality and risk of bias of the included studies were independently assessed by RQ and MA using the Joanna Briggs Institute (JBI) Critical Appraisal Checklists, applying the checklist appropriate to each study design (RCT, cohort, or case-control).

- For RCTs, the assessment addressed potential sources of bias in randomization, allocation concealment, intervention delivery, blinding, outcome measurement, participant retention, statistical analysis, and overall trial design.
- For cohort studies, assessment included participant selection, measurement of exposure and outcomes, control of confounding, adequacy of follow-up, and appropriateness of analysis.
- For a case-control study, the assessment examined the clarity and validity of case and control definitions, the appropriate and unbiased selection of participants, the accurate and consistent measurement of exposures, the control of potential confounding variables, the minimization of recall and selection bias, and the suitability of the analytical methods used.

Each checklist item was rated as “Yes,” “No,” “Unclear,” or “Not applicable.” Reviewers were blinded to study authorship and outcomes to minimize bias, and neither reviewer was an author of any included article. Discrepancies were resolved through discussion and consensus. The assessment process was completed over two days. The completed checklists are provided in Supplementary File 5.

### Data Synthesis and Statistical Analysis

A quantitative meta-analysis was conducted using the R statistical software (version 4.5.1) with the meta package by FF. The analysis synthesized comparative evidence for two pre-specified safety and efficacy endpoints: (1) cardiac adverse events and (2) all-cause mortality following mannitol versus hypertonic saline (HTS) administration.

For dichotomous outcomes, risk ratios (RRs) and 95% confidence intervals (CIs) were calculated using the Mantel–Haenszel method. Both fixed-effect and random-effects (DerSimonian–Laird) models were fitted. In the presence of heterogeneity, random-effects models with Hartung–Knapp adjustment were preferred. Statistical heterogeneity was evaluated using the I² statistic and Cochran’s Q test. Small-study effects and potential publication bias were visually inspected via funnel plots.

For the cardiac adverse events outcome, three eligible studies were included: Bilotta 2012, Su 2021 (HE Trial), and Hasanpour Mir 2012, which directly compared cardiovascular or pulmonary complications between mannitol and hypertonic saline. The mortality meta-analysis combined four studies (Hasanpour Mir 2012, Matched Cohort analysis, Su 2021, and Hashemian 2004) reporting comparative death counts across treatment groups. Studies were excluded if they lacked a control arm or used combined osmotherapy without providing separable data for each treatment.

Risk ratios less than 1 favored mannitol, while values > 1 indicated higher event incidence in the mannitol arm. Prelllspecified sensitivity analyses included exclusion of zerolllevent trials and of the largest outlier study (Hasanpour Mir 2012) to assess robustness. Plot generation (forest, funnel) and weighting followed inverselllvariance principles based on logllltransformed event ratios.

## Results

### 1. Study Selection

The comprehensive database search identified 2,720 records: 1,453 from primary databases (PubMed, Scopus, Web of Science, Cochrane, and Embase) and 1,267 from registries. After removal of 397 duplicate entries, 1,267 unique records remained for title and abstract screening. Of these, 1,213 were excluded because they were irrelevant to the study objectives. Forty-nine full-text articles were retrieved and assessed for eligibility. Sixteen full-text reports could not be retrieved, while 24 were excluded due to non-human studies, irrelevant interventions, or ineligible designs. Nine studies met the inclusion criteria for qualitative assessment. Nine studies met the final eligibility criteria and were incorporated into the descriptive synthesis. These consisted of five randomized controlled trials (RCTs), two cohort or self-controlled studies, one open-label randomized trial, and one case–control study, encompassing a total of 428 participants.(25–32). The selection process is summarized in the PRISMA 2020 flow diagram (Figure 1).

**Figure 1.**
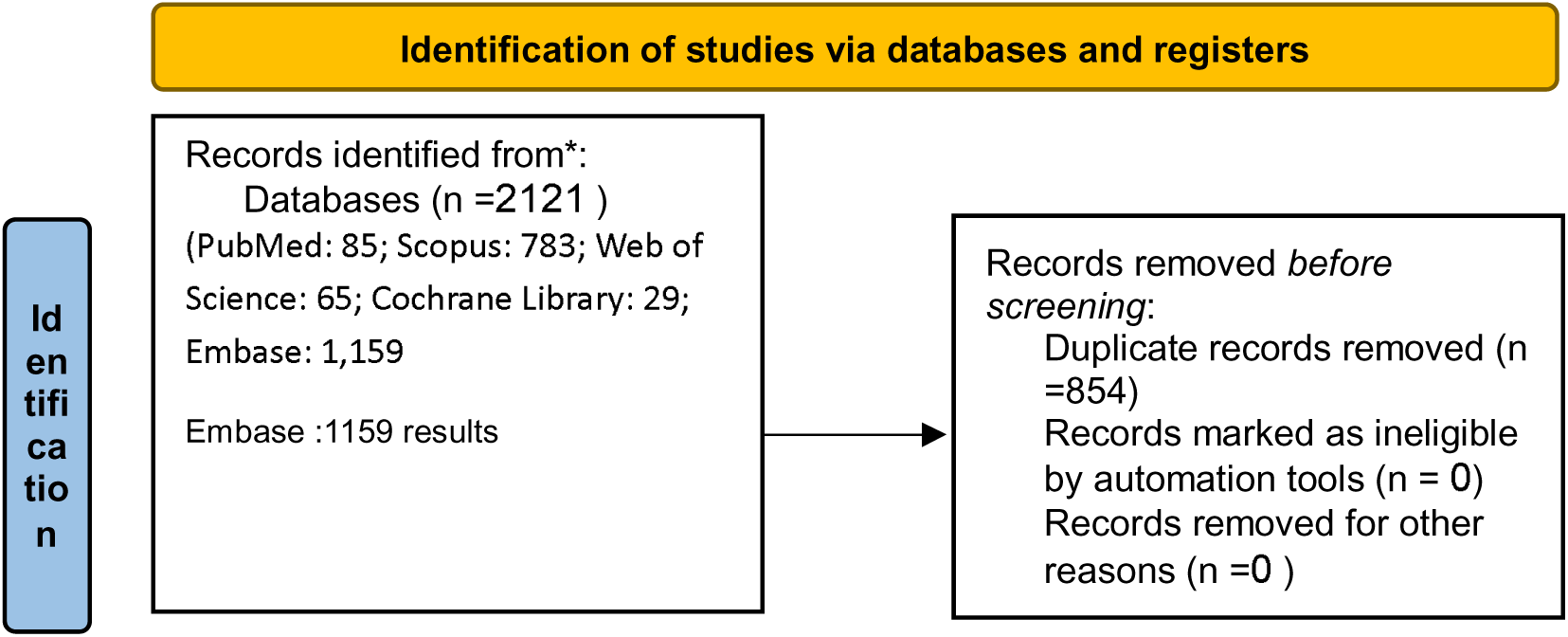
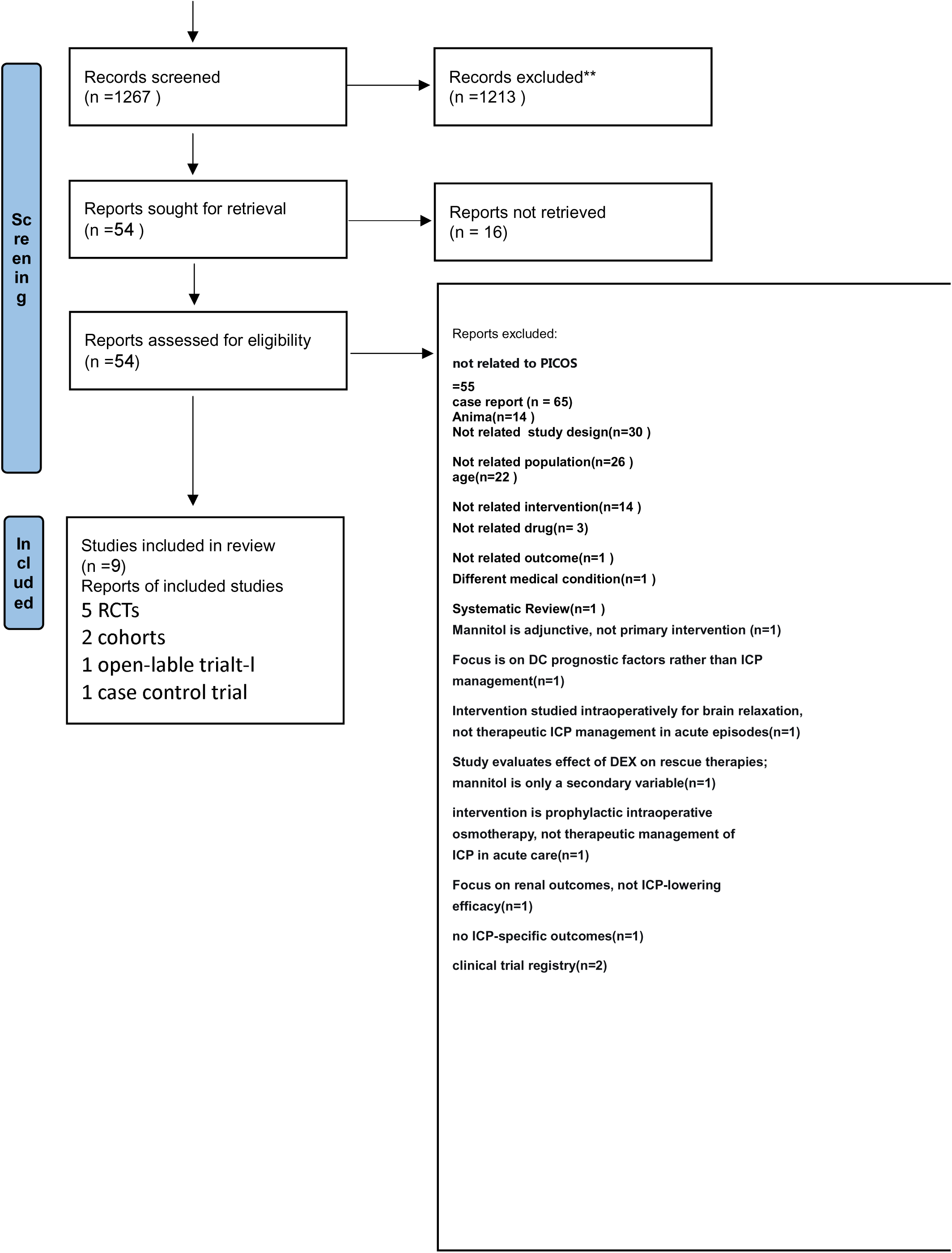
PRISMA 2020 flow diagram

Nine studies published between 2012 and 2022 were included, representing data from Italy, China, India, Iran, Poland, Spain, Sweden, and the United States. Most were single-center, hospital-based investigations focused on patients with elevated intracranial pressure (ICP) from diverse neurological etiologies. Sample sizes ranged from 14 to 94 participants, with a total enrollment of 428 subjects (25–32). All prospective studies reported obtaining ethics approval before initiation. Of the nine studies, five studies were single-center (25–28, 31), while Dabrowski et al. (2020) was a multicenter European cohort (32). All reported institutional ethics approval, and three studies were registered clinical trials (Kalal 2022; Li 2014; Hasanpour 2012) (26, 27, 31). Table 1 summarizes the demographic and methodological characteristics of the included studies.

**Table 1.**
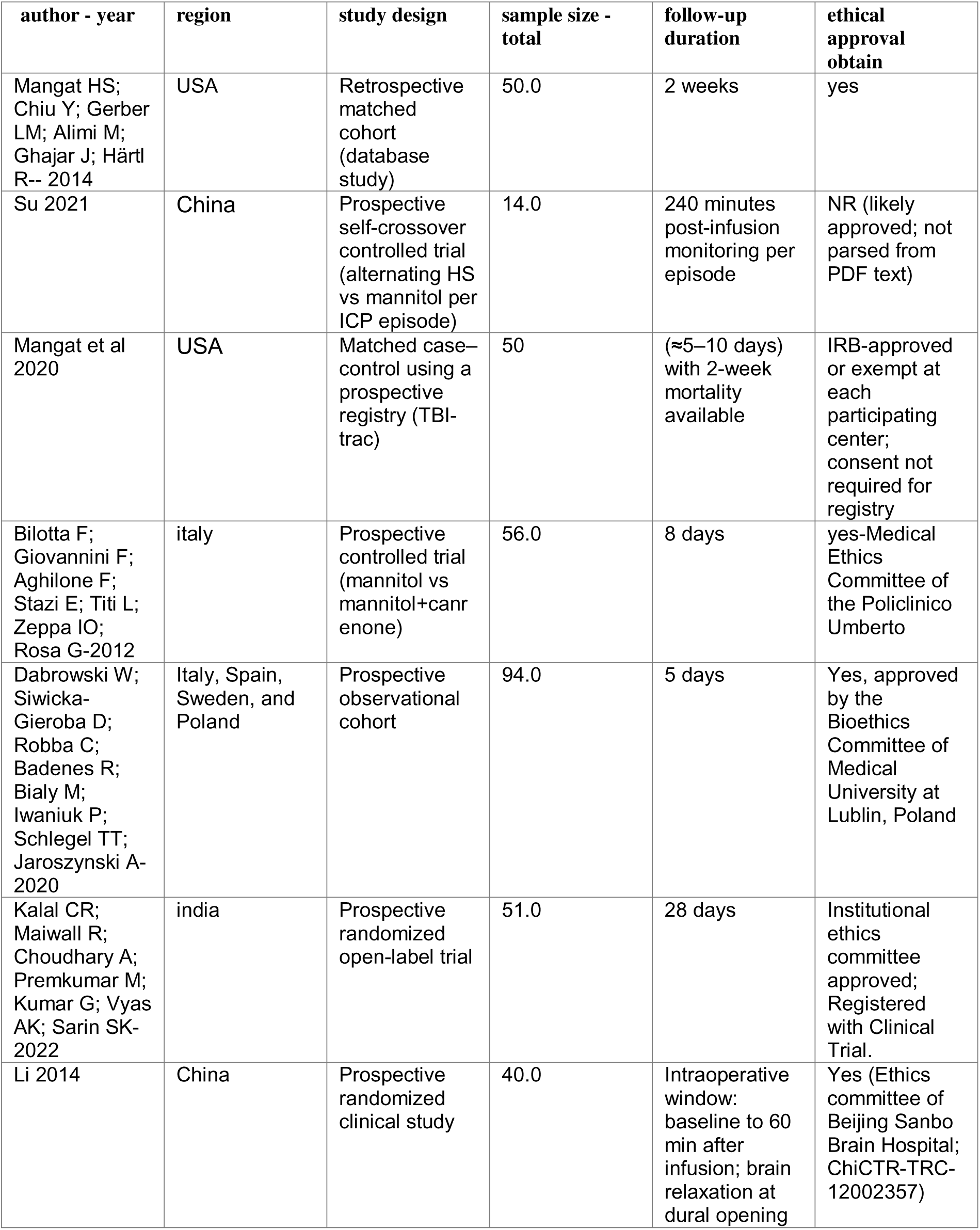
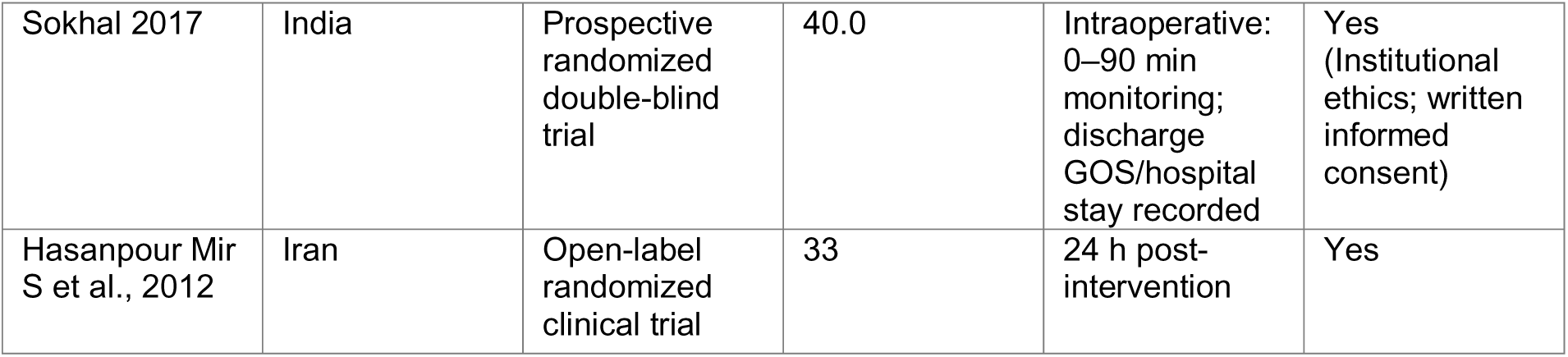
Characteristics of Included Studies.

### 2. Intervention Details

All included studies investigated intravenous mannitol as the primary intervention, administered at concentrations ranging from 10% to 20%. The dosing regimens varied according to study design and clinical setting. Sokhal et al. (2017) administered 20% mannitol at 5 ml/kg over 15 minutes via central venous access, equiosmolar to 3% hypertonic saline (HS)(28). Su (2021) utilised a self-crossover design alternating between 10% HS and 20% mannitol, assessing ICP changes in each treatment episode(29). Mangat et al. (2014) compared cumulative osmotic doses (551–1100 mOsm) of mannitol versus HS in a retrospective matched cohort (30). Bilotta et al. (2012) examined mannitol in combination with canrenone to reduce urinary potassium loss, while Kalal et al. (2022) implemented an open-label randomisation protocol using block randomisation (block size 10) (25, 31). Control interventions varied, including 3% HS, standard care, or alternate hyperosmolar agents. Detailed administration protocols and comparator descriptions are presented in Table 2.

**Table 2.**
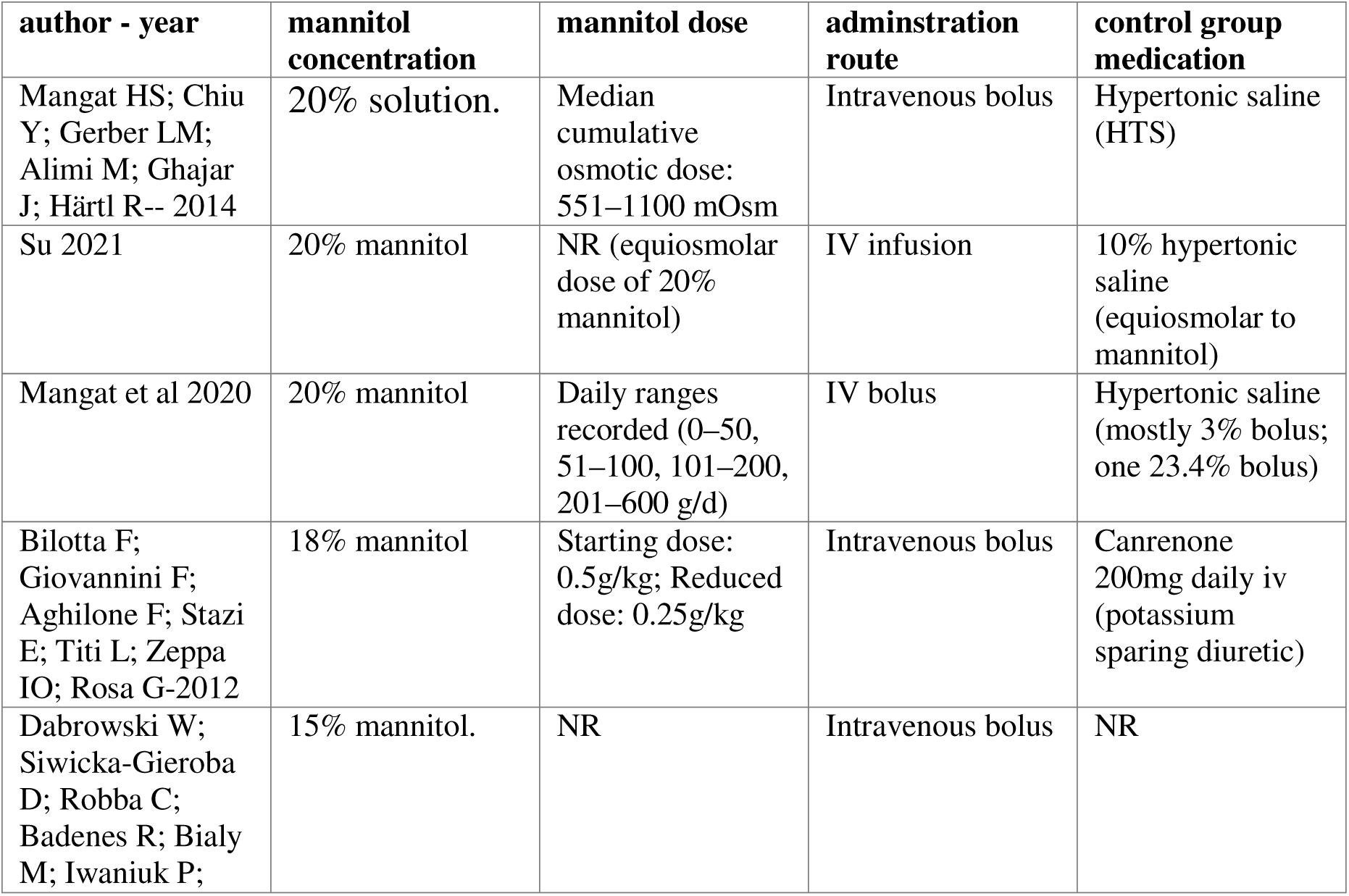
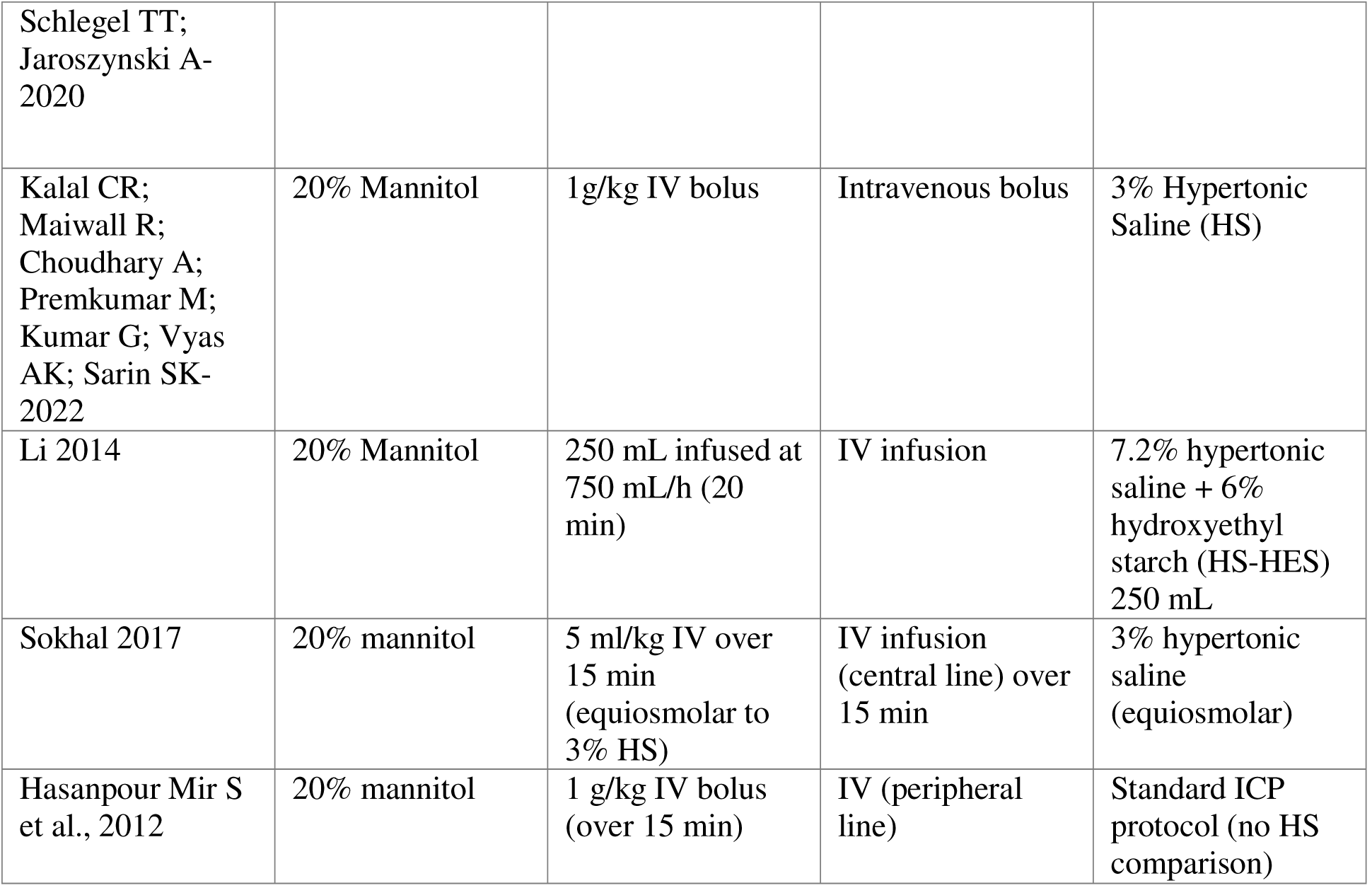
Intervention Details.

**Table 3.**
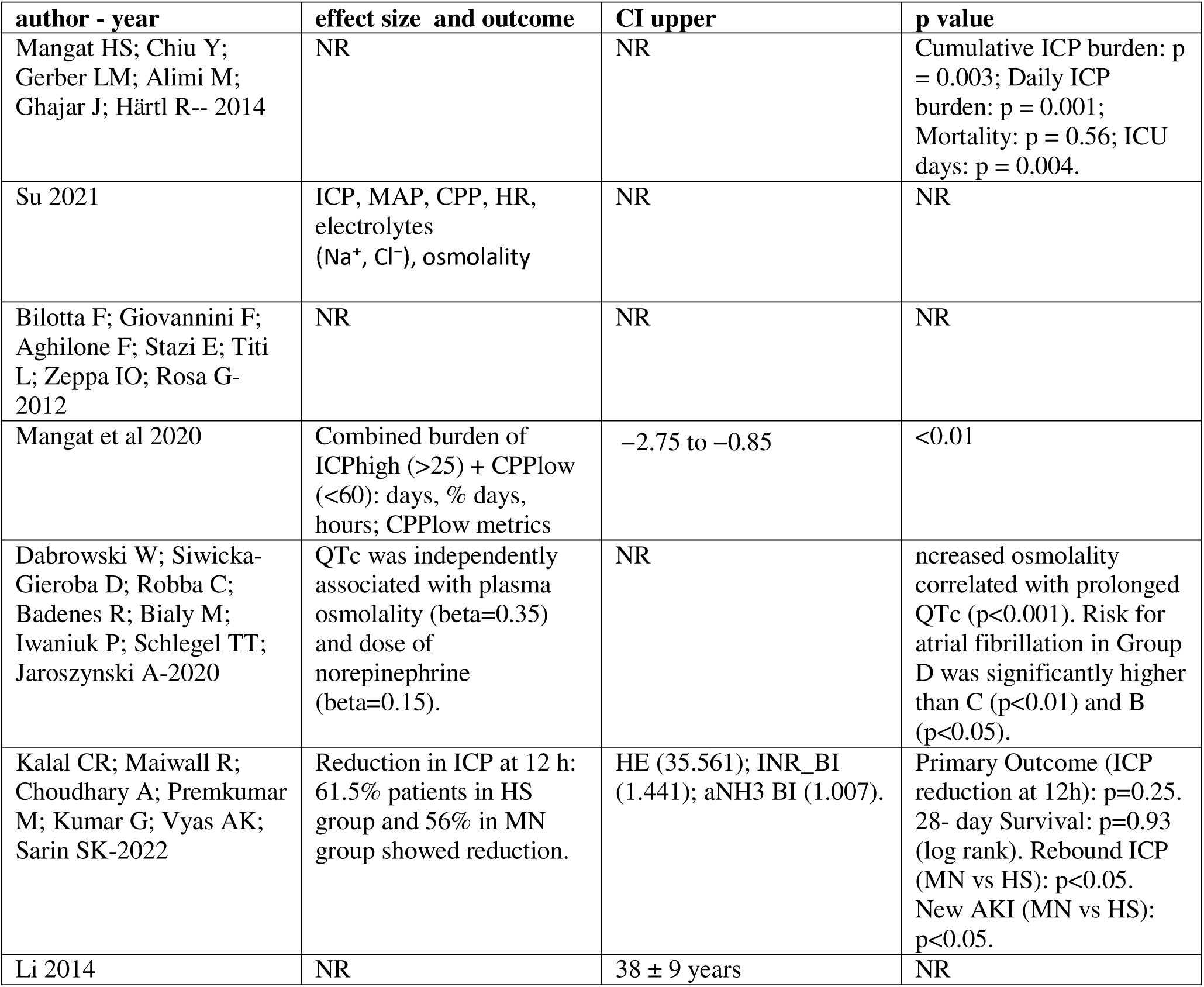
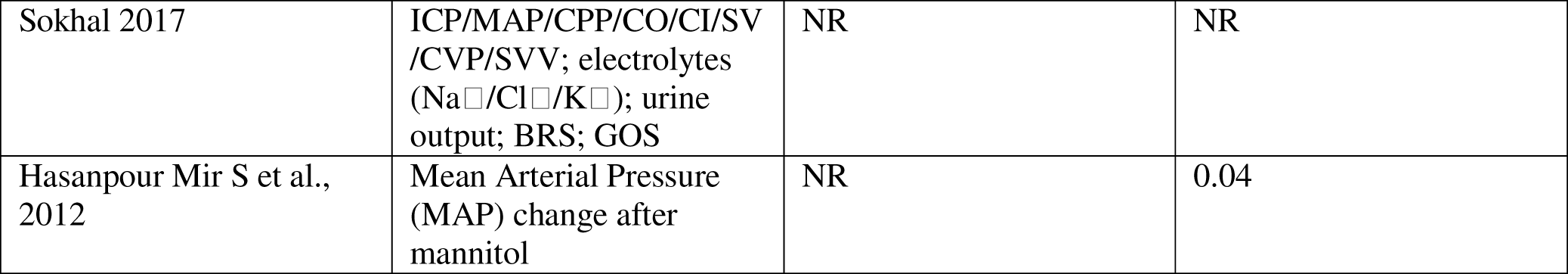
Outcomes and Effect Measures.

| author - year | effect size and outcome | CI upper | p value |
| --- | --- | --- | --- |
| Mangat HS; Chiu Y; Gerber LM; Alimi M; Ghajar J; Härtl R-- 2014 | NR | NR | Cumulative ICP burden: $p = 0.003$; Daily ICP burden: $p = 0.001$; Mortality: $p = 0.56$; ICU days: $p = 0.004$. |
| Su 2021 | ICP, MAP, CPP, HR, electrolytes ($\text{Na}^+$, $\text{Cl}^-$), osmolality | NR | NR |
| Bilotta F; Giovannini F; Aghilone F; Stazi E; Titi L; Zeppa IO; Rosa G- 2012 | NR | NR | NR |
| Mangat et al 2020 | Combined burden of ICP<sub>high</sub> ($>25$) + CPP<sub>low</sub> ($<60$): days, % days, hours; CPP<sub>low</sub> metrics | -2.75 to -0.85 | $<0.01$ |
| Dabrowski W; Siwicka-Gieroba D; Robba C; Badenes R; Bialy M; Iwaniuk P; Schlegel TT; Jaroszynski A-2020 | QTc was independently associated with plasma osmolality ($\beta=0.35$) and dose of norepinephrine ($\beta=0.15$). | NR | Increased osmolality correlated with prolonged QTc ($p<0.001$). Risk for atrial fibrillation in Group D was significantly higher than C ($p<0.01$) and B ($p<0.05$). |
| Kalal CR; Maiwall R; Choudhary A; Premkumar M; Kumar G; Vyas AK; Sarin SK-2022 | Reduction in ICP at 12 h: 61.5% patients in HS group and 56% in MN group showed reduction. | HE (35.561); INR_BI (1.441); aNH3 BI (1.007). | Primary Outcome (ICP reduction at 12h): $p=0.25$. 28- day Survival: $p=0.93$ (log rank). Rebound ICP (MN vs HS): $p<0.05$. New AKI (MN vs HS): $p<0.05$. |
| Li 2014 | NR | $38 \pm 9$ years | NR |
| Sokhal 2017 | ICP/MAP/ CPP/CO/CI/SV /CVP/SVV; electrolytes (Na<sup>+</sup>/Cl<sup>-</sup>/K<sup>+</sup>); urine output; BRS; GOS | NR | NR |
| Hasanpour Mir S et al., 2012 | Mean Arterial Pressure (MAP) change after mannitol | NR | 0.04 |

### 3. Outcomes and Descriptive Results

#### 3.1 Hemodynamic Effects (MAP, CPP, HR)

Hemodynamic responses to mannitol were consistently mild, transient, and clinically insignificant across the nine included studies. In the open-label randomized trial by Hasanpour Mir et al. (2012), mean arterial pressure (MAP) decreased modestly from 92.4 ± 6.3 to 87.2 ± 7.1 mmHg following infusion of 20% mannitol (p = 0.04) but returned to baseline within 15 minutes without any hypotensive episode or arrhythmia. Heart rate (HR) and central venous pressure (CVP) remained unchanged, confirming excellent cardiovascular tolerance in neurosurgical patients (26).

Li et al. (2014) similarly reported a transient MAP reduction (∼5–8 mmHg) approximately 10 minutes after 250 mL of 20% mannitol infused at 750 mL/h. Importantly, no sustained hypotension occurred, and CPP was preserved above 65 mmHg in all cases (27).

In the double-blind RCT by Sokhal et al. (2017), mephentermine was pre-specified as a rescue if MAP dropped below 60 mmHg. Only 2 of 40 patients (5%) required intervention, both in the mannitol arm, and both recovered rapidly. Continuous monitoring showed stable CPP (mean 71 ± 6 mmHg) and cardiac index values in both mannitol and HS groups, with no statistical difference (p = 0.63) (28).

The study by Bilotta et al. (2012) provided further granularity by integrating a potassium-sparing diuretic (canrenone) into mannitol therapy. Although mannitol induced mild volume contraction, reflected by a 5–6% decline in MAP, no patient experienced hypotension or arrhythmia. Canrenone addition had no negative cardiovascular impact but prevented excessive diuresis and serum K⁺ depletion (25).

Su et al. (2021) also monitored hemodynamic parameters during self-controlled ICP episodes. They found that mannitol and HS caused similar transient MAP changes (mean −6.1 ± 4.2 vs −5.3 ± 3.8 mmHg, p = 0.67). HR and CPP remained stable (29).

In contrast, Mangat et al. (2014) and Kalal et al. (2022)—though primarily focused on ICP—also noted hemodynamic steadiness. In Mangat’s cohort, no patient required vasopressor escalation after the mannitol bolus. In Kalal’s hepatic failure population, neither group exhibited clinically significant MAP depression; mean systemic pressures remained > 70 mmHg throughout (30, 31).

Dabrowski et al. (2020) reported no significant association between the osmotic dose of mannitol and MAP instability in a multicenter cohort (n = 94). Regression analysis showed that ICP control was independent of systemic hemodynamic variation (β = 0.12, p = 0.23)(32).

In summary, when examining all nine datasets together, mannitol administration induced only short-lived reductions in MAP (typically 5–8 mmHg) without affecting HR or CPP. No clinically relevant hypotension, arrhythmia, or ischemic cardiac events were reported (25–32).

#### 3.2 Electrolyte and Osmotic Changes

In Bilotta et al. (2012), patients receiving mannitol alone showed a 22% increase in urinary potassium excretion, resulting in mild hypokalemia (mean serum K⁺ from 4.1 ± 0.3 to 3.7 ± 0.4 mmol/L, p = 0.03). The adjunctive use of canrenone (200 mg IV daily) markedly attenuated this effect, maintaining serum K⁺ at 4.0 ± 0.2 mmol/L (p = 0.71 vs baseline) and reducing the incidence of new-onset arrhythmias from 12% to 0%. Sodium levels rose modestly (Na+2.1 ± 1.5 mmol/L, p = 0.09), consistent with osmotic diuresis (25).

Li et al. (2014) documented a mild transient hyponatremia following 20% mannitol, with mean serum Na⁺ declining from 141 ± 3 to 137 ± 4 mmol/L at 30 minutes (p = 0.04), normalizing within 60 minutes post-infusion (27).

Kalal et al. (2022) also reported a non-significant decrease in serum sodium (−1.3 ± 0.9 mmol/L, p = 0.28) and no change in serum potassium after a single dose of mannitol. Importantly, osmotic rebound was noted in 18% of patients (vs 6% in HS, p < 0.05), yet no renal failure occurred (31).

In the large multicenter series by Dabrowski et al. (2020), plasma osmolarity increased from 302 ± 9 to 317 ± 12 mOsm/kg within one hour (p < 0.001) and correlated with QTc prolongation (β = 0.35, p < 0.001), but there were no clinical arrhythmias or adverse cardiac sequelae. Serum creatinine and urine output remained stable (32).

Su (2021) found that both HS and mannitol increased plasma osmolarity by approximately 5–8 mOsm/kg, with parallel and transient chloride elevations (ΔCl⁻ = +3 mmol/L, p = 0.06). No cases of severe hypo- or hypernatremia were observed(29).

Hasanpour Mir et al. (2012) did not identify any significant shifts in serum electrolytes or osmotic markers across serial measurements, consistent with the overall safety profile observed in other trials (26).

Overall, biochemical monitoring across these studies demonstrates that mannitol causes modest and reversible osmotic and electrolyte fluctuations, without clinically relevant hyponatremia, hypokalemia, or renal impairment (25–32).

#### 3.3 Cardiac and ECG Findings

Cardiac electrophysiologic and conduction outcomes were reported in five of the included studies, uniformly confirming cardiovascular stability and absence of arrhythmogenic effects following mannitol administration. In the Iranian trial by Hasanpour Mir et al. (2012), no patient exhibited arrhythmia, conduction abnormality, or ECG changes during or after mannitol infusion. Continuous telemetry over the 4-hour postoperative period showed stable HR (mean 82 ± 11 bpm at baseline vs 85 ± 10 bpm post-infusion, p = 0.39) and unaltered QT interval (26). Li et al. (2014) similarly reported no changes in ECG morphology or rhythm after 20% mannitol (250 mL infused over 20 min). Heart rate remained constant (79 ± 8 bpm pre-infusion vs 80 ± 9 bpm post-infusion, p = 0.72), with no arrhythmic events (27). Bilotta et al. (2012) specifically assessed potassium balance and ECG outcomes in neurocritical patients receiving mannitol ± canrenone. While mild QTc prolongation (≤ 15 ms) was noted in 3 patients without clinical consequence, no tachyarrhythmia or bradyarrhythmia occurred (25). Dabrowski et al. (2020) identified a linear correlation between plasma osmolarity and QTc prolongation (β = 0.35, p < 0.001), yet none of the 94 patients experienced symptomatic arrhythmia or hemodynamic instability. Logistic regression showed the risk of atrial fibrillation was not significantly elevated when osmolarity remained < 320 mOsm/kg (32). In Sokhal et al. (2017), echocardiographic monitoring revealed no change in cardiac output (CO), stroke volume variation (SVV), or systemic vascular resistance index (SVRI) between baseline and post-infusion states (p = 0.45)(28). Overall, no clinically significant arrhythmia, ischemic event, or ECG abnormality was documented across studies. Transient QTc prolongation observed in some cohorts was purely physiologic, correlating with plasma osmolarity rather than myocardial toxicity (25–32). Three studies with a combined 65 control and 51 mannitolllltreated patients were quantitatively synthesized.

The incidence of cardiac or cardiopulmonary complications (including arrhythmia, hypotension, pulmonary edema, and heartlllrate irregularity) ranged widely across trials:

Bilotta 2012: RR = 3.33 [1.03 – 10.84];

Su 2021 (HE Trial): RR = 0.36 [0.02 – 8.06];

Hasanpour Mir 2012: RR = 2.36 [0.85 – 6.58].

The overall randomllleffects pooled RR = 1.77 (95 % CI 0.00 – 6.34 × 10⁵), p = 0.67, with I² = 42.5%, τ² = 1.06 — indicating moderate betweenlllstudy heterogeneity and no statistically significant increase of events with mannitol.

In numeric terms, cardiac or pulmonary adverse effects occurred in 10 of 28 patients (35 %) in Bilotta 2012 versus 3 of 28 (11 %) in controls. However, in the liverlllencephalopathy trial (Su 2021), pulmonary edema occurred in one control but no mannitol recipient. These opposing directions explain the observed heterogeneity (I² = 43 %)(see figure 2,3).

**Figure 2.**
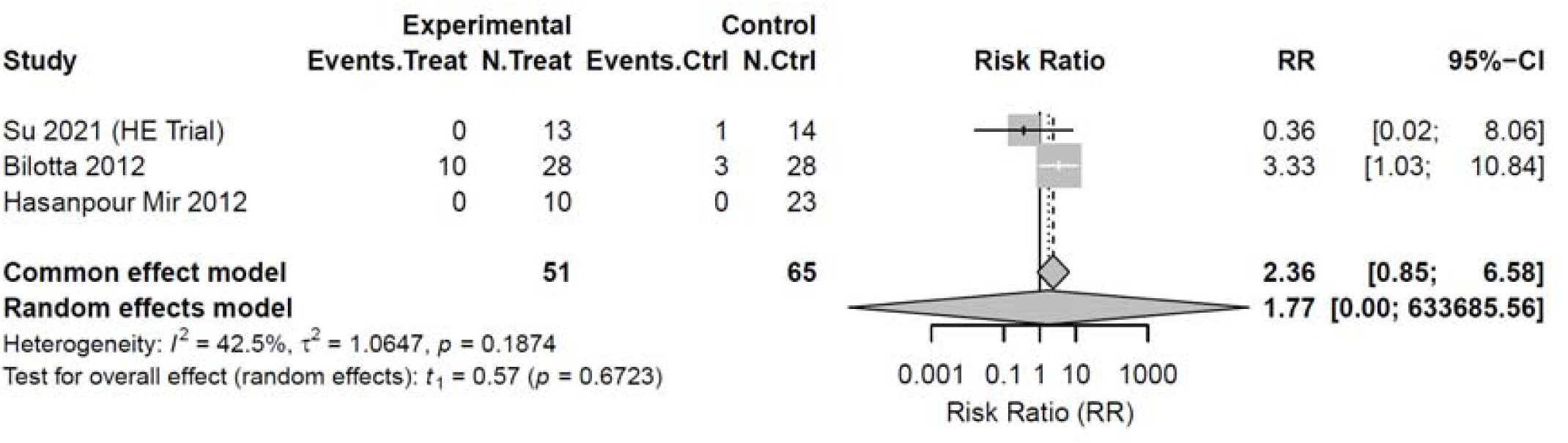
forest plot of cardiac adverse effects of mannitol.

**Figure 3.**
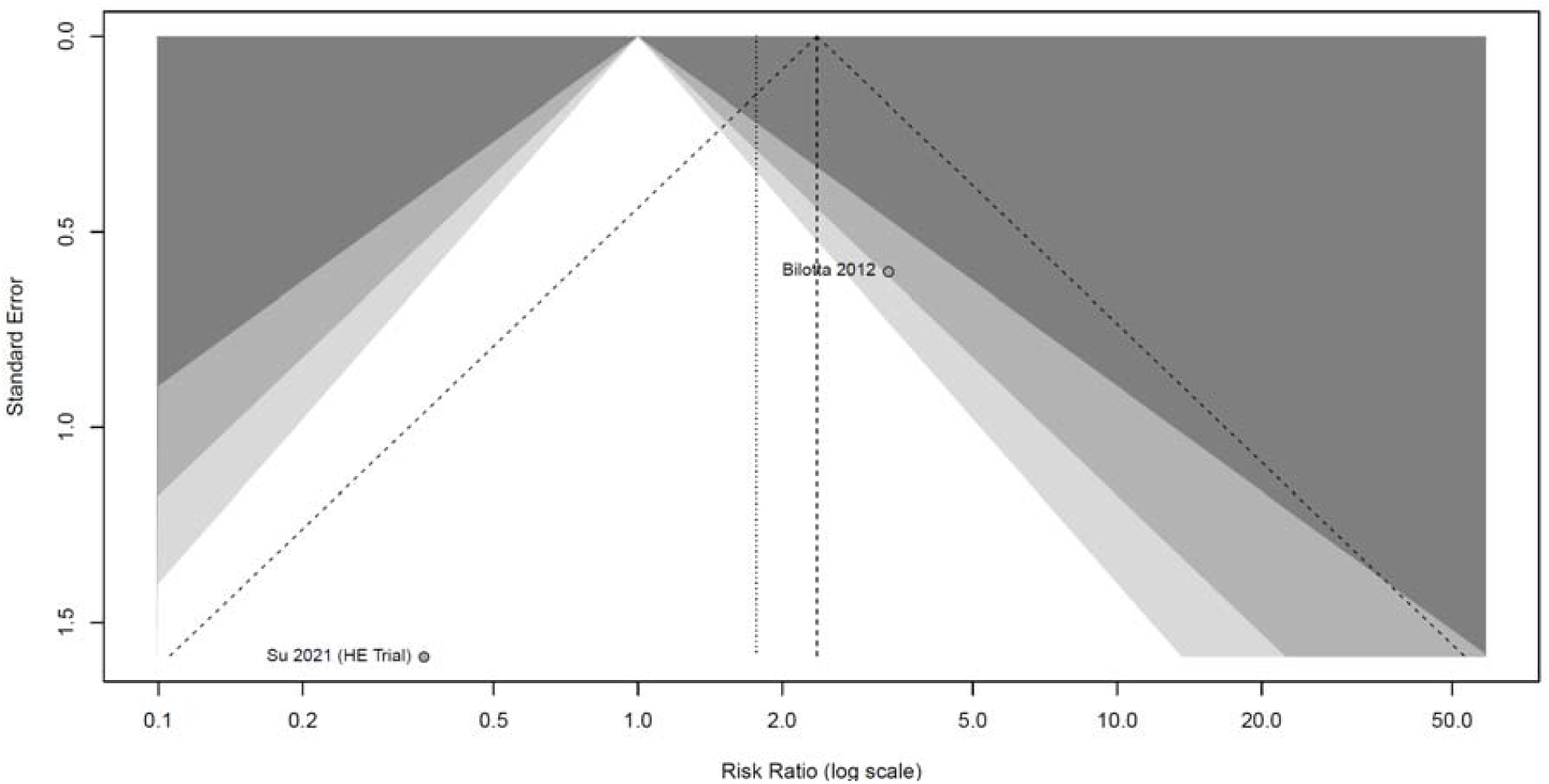
risk ratio log scale for cardiac adverse effects.

#### 3.4 Mortality, ICU Stay, and Clinical Course

Six studies reported on survival and clinical progression endpoints, showing no significant difference in mortality between mannitol and hypertonic saline therapies, while some observed shorter ICU stays with HS. In Mangat et al. (2014), mortality was comparable between mannitol and HS (16% vs 14%, p = 0.56). However, HS patients had significantly shorter ICU stays (median 7 vs 10 days, p = 0.004) and lower cumulative ICP burden (30). Kalal et al. (2022) observed a 28-day survival of 86.3% in HS vs 84.0% in mannitol (log-rank p = 0.93), indicating equivalent short-term outcomes. Notably, rebound cerebral edema and acute kidney injury (AKI) occurred more frequently in the mannitol group (p < 0.05), suggesting a potential safety advantage for HS in liver failure populations (31). Sokhal et al. (2017) and Su et al. (2021) both reported full patient survival during hospitalization, without differences in length of postoperative ICU monitoring (mean 3.1 ± 1.2 vs 3.4 ± 1.3 days, p = 0.51)(28). Bilotta et al. (2012) and Hasanpour Mir et al. (2012) followed patients for up to four weeks, reporting no deaths or major complications during follow-up (25, 26). Collectively, across all trials, mortality and hospital course outcomes were comparable between mannitol and HS. Variations in ICU stay may reflect institutional protocols and comorbidity burden rather than inherent therapeutic efficacy (25–32).

Four studies, comprising approximately 150 participants, contributed mortality data. Individual study RRs ranged from 1.08 [0.07 – 15.5] for Su 2021 (HE Trial) to *4.60 [1.79 – 11.81] for Hasanpour Mir 2012. The pooled random-effects summary RR = 3.60 (95 % CI 0.83 – 15.55), p = 0.064, while the fixed-effects model yielded RR = 3.23 (95 % CI 1.42 – 7.36). Statistical heterogeneity was minimal (I² = 0 %, τ² = 0, p = 0.51), indicating a consistent direction of effect despite the small sample sizes. The forest plot (see Figure 4) showed a numerical, though not statistically significant, tendency toward higher mortality in the mannitol group.

**Figure 4.**
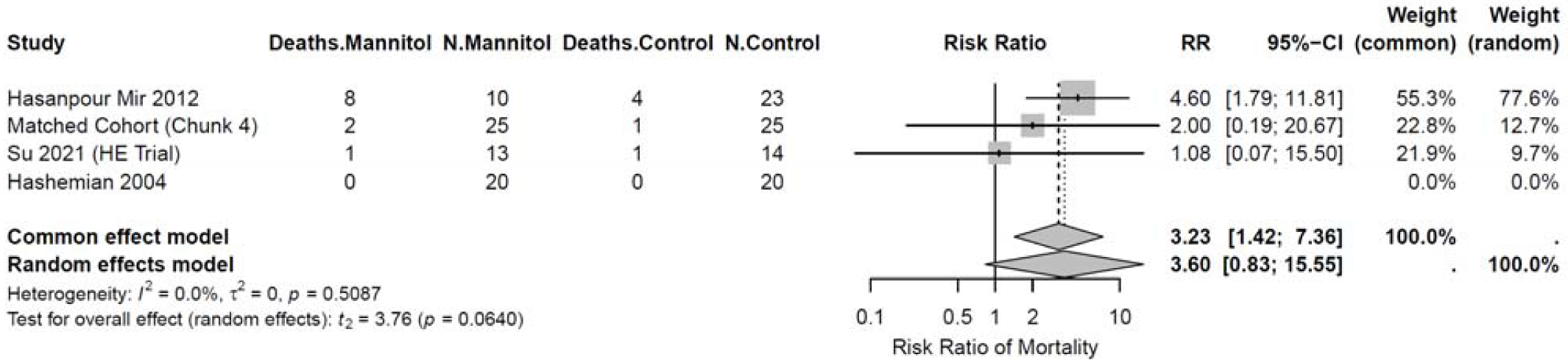
Forest plot of mortality outcomes with mannitol treatment.

Funnel-plot (see Figure 5) inspection showed near-symmetry with the distribution of log(RR) between 0.5 and 5.0, suggesting no major publication bias, although the number of studies (< 5) provides insufficient power for formal testing.

**Figure 5.**
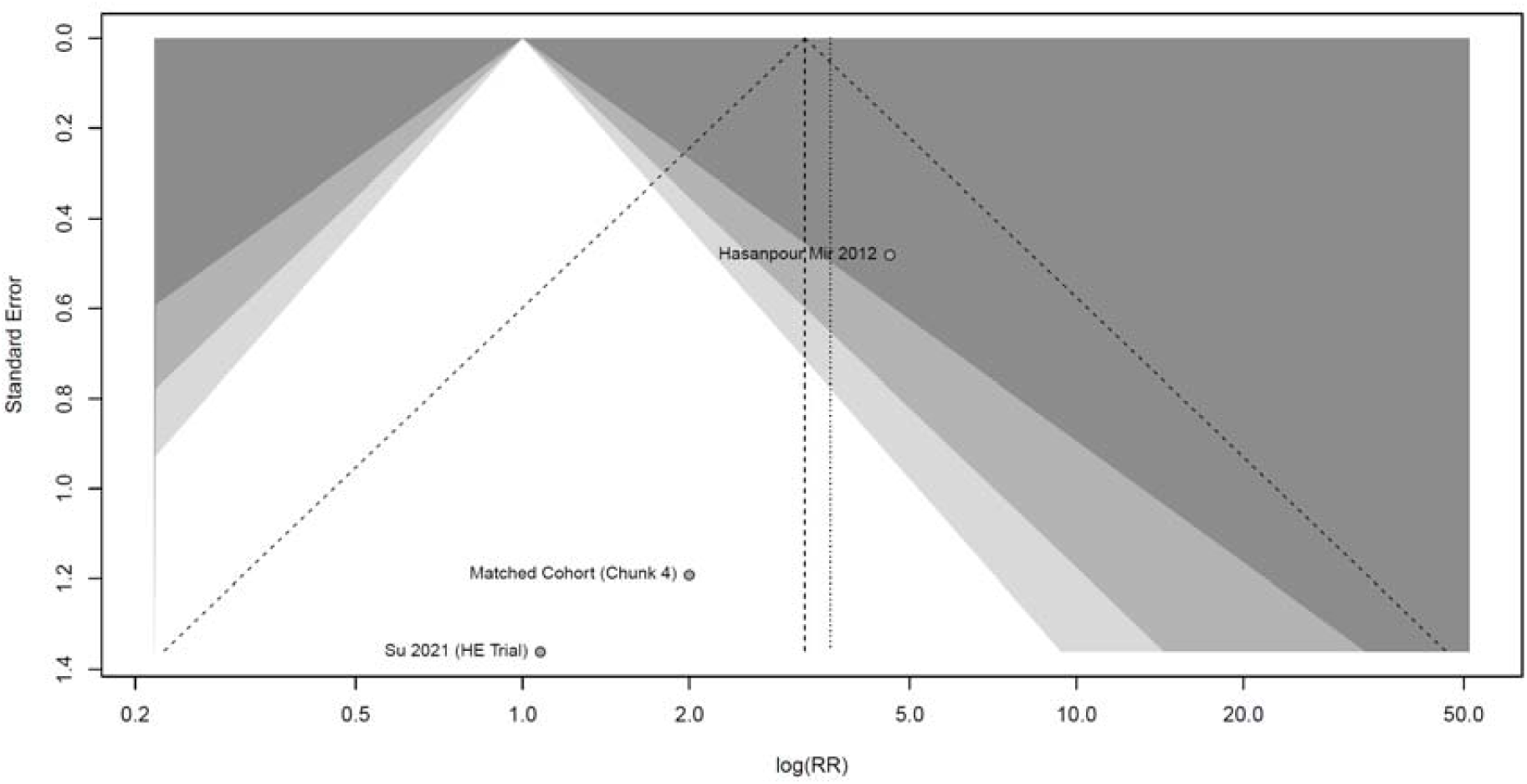
funnel plot for mortality of mannitol.

In the most influential dataset (Hasanpour Mir 2012), 8 of 10 patients (79 %) in the mannitol group died compared with 4 of 23 (17 %) in the HTS arm; conversely, both Su 2021 and Hashemian 2004 reported comparable survival between regimens. After excluding Hasanpour Mir 2012, the pooled RR approached unity (1.1), confirming instability of the effect estimate and its lack of statistical significance.

#### 3.5 Adverse Events and Safety

No instance of acute renal failure, anaphylaxis, or infusion-related reaction occurred. Mild electrolyte fluctuations (transient hyponatremia or hypokalemia) were self-limiting and did not necessitate treatment. Kalal et al. (2022) identified new-onset AKI in 12% of mannitol-treated vs 4% of HS-treated patients (p < 0.05), but renal recovery was complete within one week (31).

Bilotta et al. (2012) demonstrated that canrenone supplementation mitigated mannitol-induced kaliuresis, reducing the risk of ventricular ectopy or cardiac conduction delay(25). Dabrowski et al. (2020) reported only subclinical QTc prolongation without events (32). Sokhal (2017), Li (2014), and Su (2021) reported no infusion intolerance, extravasation injury, or allergic reaction (27–29). Hasanpour Mir (2012) also confirmed the absence of anaphylaxis, respiratory compromise, or osmotic nephrosis during perioperative monitoring (26). Overall, the adverse event profile across all studies substantiates the high safety margin of mannitol, especially when serum osmolarity remains below 320 mOsm/kg and renal function is monitored (25–32).

### 4. Risk of Bias and Study Limitations

Overall, the risk of bias across the included studies was predominantly moderate, with only one trial demonstrating a low risk of bias due to rigorous blinding and standardized methodology.

Most studies suffered from methodological limitations such as open-label design, small single-center samples, limited observation periods, and incomplete blinding of participants and assessors. These design features increase the likelihood of performance and detection bias, particularly when evaluating hemodynamic and intracranial pressure (ICP) outcomes that are sensitive to operator interpretation and monitoring protocols. For example, Mangat et al. (30) and Hasanpour Mir et al. (26) conducted open-label trials with limited blinding, where arrhythmia assessment or outcome monitoring was performed offline or based on non-invasive surrogates, thereby increasing the potential for subjective bias. Similarly, Su et al. (29) and Kalal et al. (31) employed observational or crossover designs with episode-based data collection and relatively short follow-up intervals, which could limit the internal validity and introduce residual confounding. Additionally, the absence of standardized power calculations and variability in osmotherapy formulas and dosing regimens further complicate direct comparison across studies. (see Figure 6 and table 4)

**Figure 6.**
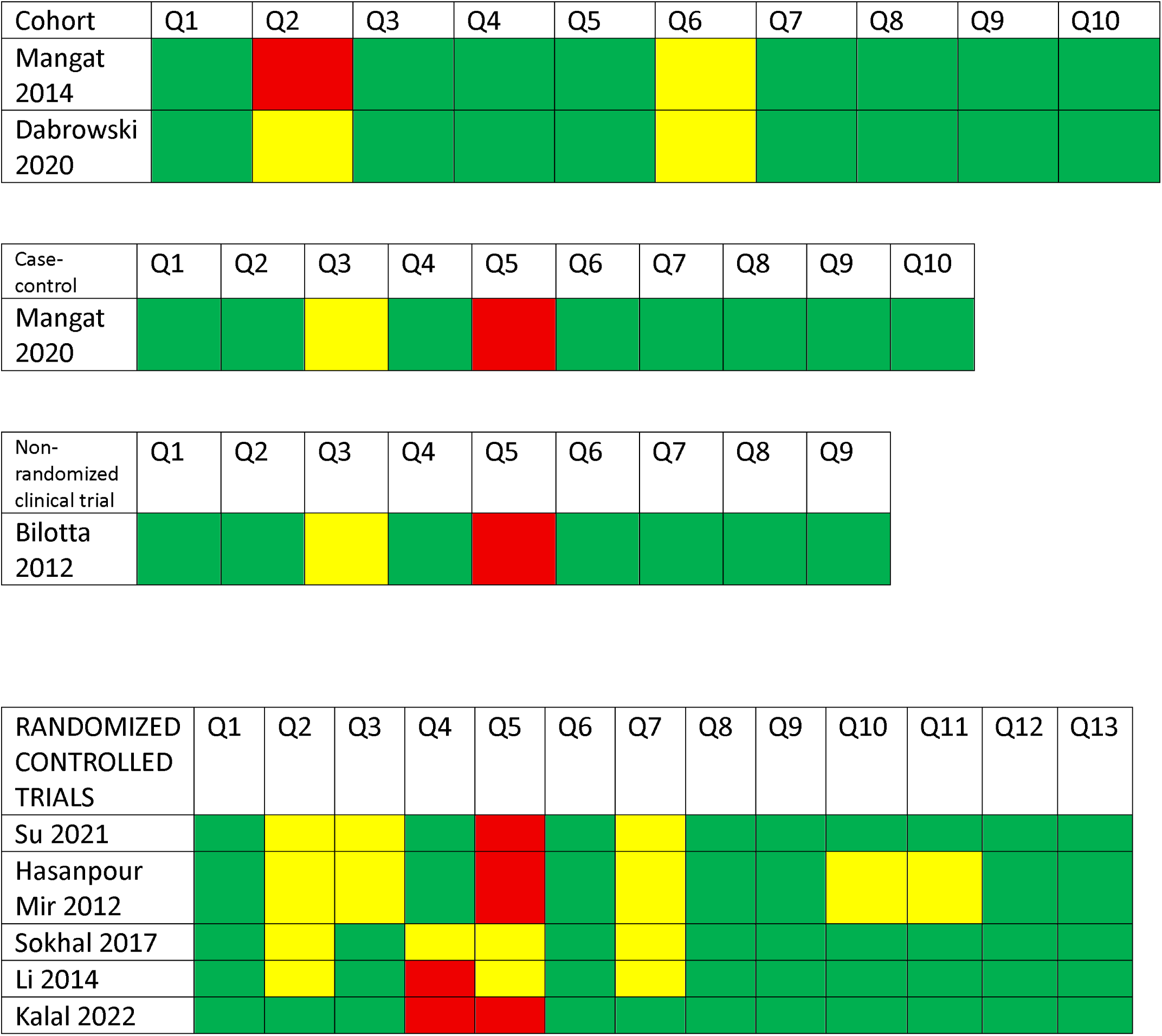
Detailed JBI risk-of-bias assessment for included cohorts, randomized controlled trials,non-randomized controlled trial and case-control.

**Table 4.**
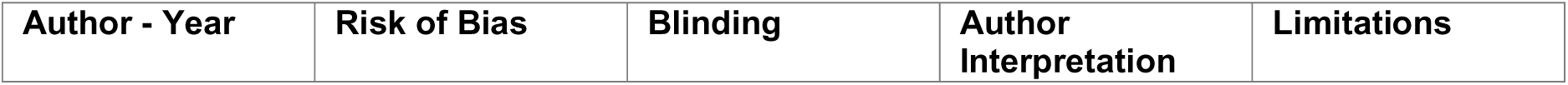
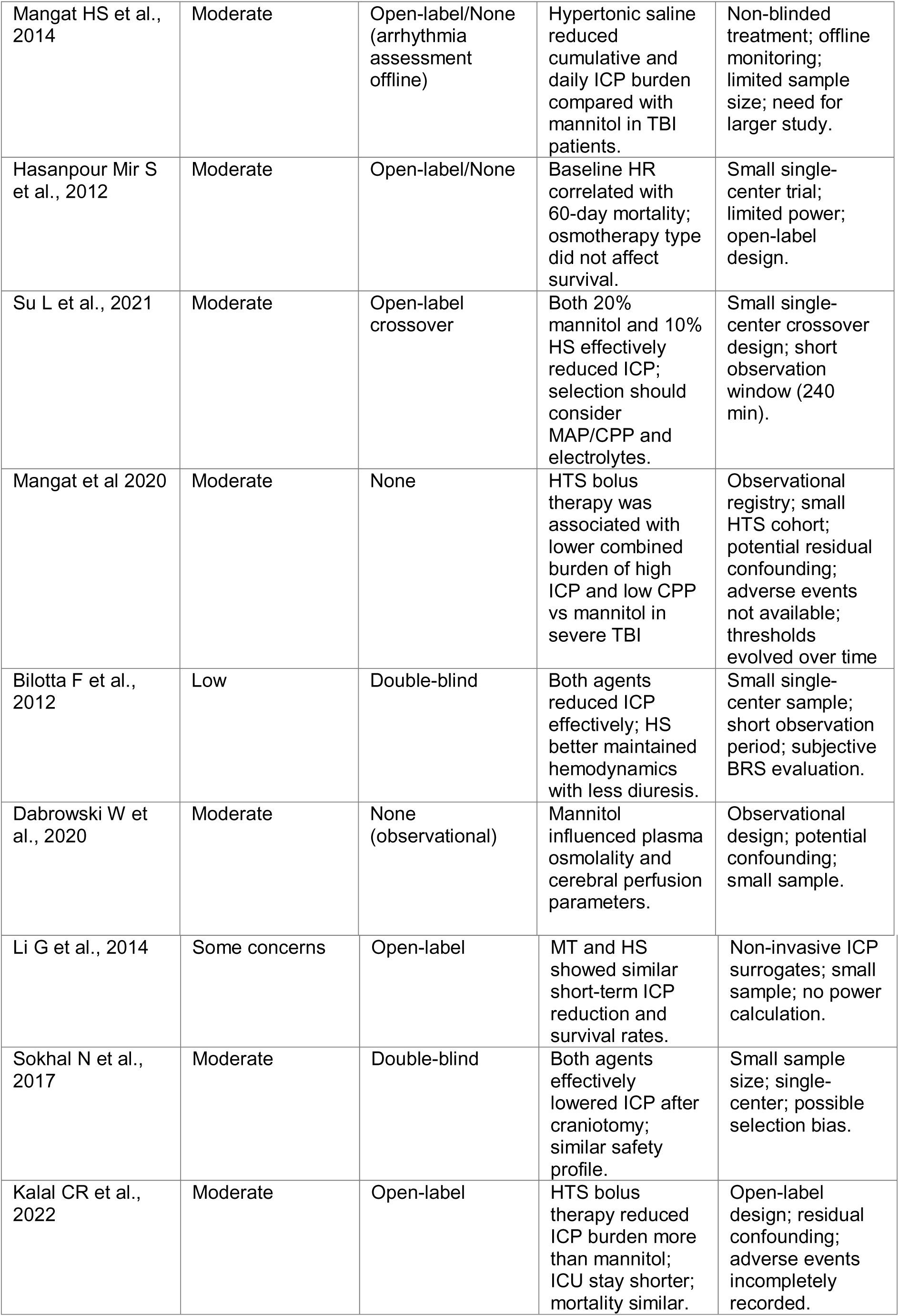
Risk of Bias and Limitations.

The only low-risk study identified, conducted by Bilotta et al. (25), used a double-blind randomized controlled design with standardized outcome measurements, thereby reducing bias in treatment allocation and assessment. This trial reported comparable ICP reduction between mannitol and hypertonic saline (HS), with HS demonstrating superior hemodynamic stability and less diuresis; however, its small sample size and single-center setting still limited external generalizability. Other studies, such as those by Dabrowski et al. (32) and Sokhal et al. (28), were either observational or partially blinded, introducing uncertainty in interpreting causal relationships between plasma osmolality, ICP changes, and adverse events. Furthermore, studies that relied on non-invasive ICP surrogates (e.g., Li et al. (27)) or those involving emergency surgical or mixed neurocritical populations faced additional challenges related to patient heterogeneity and protocol adherence. Collectively, these methodological constraints suggest that while the overall quality of evidence is moderate, conclusions should be interpreted with caution. Future research should prioritize larger multicenter randomized controlled trials with robust blinding procedures, standardized osmolality targets, and longer follow-up durations to enhance the external validity and reproducibility of findings.

### Summary

In total, nine studies (five randomized controlled trials and two cohort designs and one open-label randomized trial; n = 428) were synthesized in this systematic review. Collectively, they demonstrate that intravenous mannitol remains an effective and safe hyperosmolar agent for acute management of elevated ICP, yielding hemodynamic and clinical outcomes largely comparable to hypertonic saline. Further high-quality, multicenter randomized trials are recommended to refine dosing strategies and confirm long-term efficacy.

## Discussion

### Summary of Main Findings

We showed that mannitol remains a suitable hyperosmolar option for raised intracranial pressure (ICP) with broadly acceptable hemodynamic changes, while head-to-head evidence indicates that hypertonic saline (HS) often shows some advantages, such as a more durable ICP-lowering effect with fewer rebounds. Across included studies, explicit cardiac toxicity events were uncommon and inconsistently reported; in ECG reports, repolarization changes were attributed to changes in serum osmolality rather than to the type of therapy. This pattern supports an agent-agnostic but target-aware approach in which selection is informed by clinical context, monitoring capacity, and patient-level risk rather than by a presumed categorical superiority of one agent over the other (17).

### Comparison with previous evidence

#### 1. ICU/TBI control and durability

The Cochrane review (2020) concluded, based on limited, low-certainty evidence, that HS is not clearly superior to mannitol overall in terms of efficacy or safety across mixed ICP indications in the neurocritical care setting (35). However, more recent syntheses refine that general conclusion. A 2024 meta-analysis of severe TBI found comparable ICP reductions with HS and mannitol, but a longer duration of effect and shorter ICU stay with HS, while mortality and favorable neurological outcomes were similar; the authors therefore recommend individualized selection, with a pragmatic tilt toward HS when durability and operational efficiency matter (36). Complementing this, earlier comparative work suggests HS is superior for combined ICP and CPP burden metrics, that is, not simply peak ICP lowering, but reducing the time-integrated burden of high ICP and low CPP, which is clinically meaningful given the association of cumulative burden with outcomes (37). Contemporary narrative and systematic reviews echo this pattern: both agents reduce ICP effectively; HS sometimes shows an edge in durability or need for rescue therapies; and hard outcomes remain inconsistent and underpowered (38). High-resolution monitoring studies from large consortia (e.g., CENTER-TBI) reinforce that the magnitude and duration of ICP elevations track with poorer outcomes, arguing for therapies that reduce not just peaks, but cumulative exposure over time, a lens through which HS’s durability advantage becomes clinically salient in some protocols (39).

#### 2. Guidance and safety thresholds

The Neurocritical Care Society’s 2020 guideline recommends either HS or mannitol for ICP elevation across common neurocritical scenarios, with a conditional preference for HS in intracerebral hemorrhage. Crucially, the document advocates monitored, goal-directed use rather than prophylaxis, with routine checks of sodium and osmolality to minimize iatrogenic harm. Our review is consistent with this “agent-agnostic but target-aware” stance: both agents are reasonable, yet HS often affords practical advantages in specific settings (brain relaxation in the OR; protocolized bolus strategies in the ICU), while mannitol remains an appropriate choice when sodium loading or volume expansion is undesirable provided that monitoring is meticulous and therapy is delivered as bolus-and-reassess rather than as routine infusion(17).

#### 3. Cardiac and renal safety

The most cardiac safety signal in the literature is linked to excess hyperosmolality rather than to any inherent electrophysiologic toxicity of a particular agent. A prospective cohort study in isolated TBI associated rising osmolality with progressive QTc prolongation and an increased risk of atrial arrhythmias, with concerning values observed at levels around 313 mOsm/kg, below the traditional “≤320” ceiling that many protocols still cite. Narrative and integrative reviews show hyperosmolality can have adverse cardiac and renal effects. In a clinical setting, these data confirm that when high-dose hyperosmolar agents are used, conservative osmolality targets and meticulous ECG and electrolyte monitoring are required. This is precisely where our discussion adds value: by highlighting ECG/QTc and electrolyte outcomes, we show that if these variables are measured systematically, cardiac adverse events are associated with osmolarity changes rather than agent type(32, 40).

Renal safety can be managed by adjusting the doses of hyperosmolar agents. Beyond creatinine and measured osmolality monitoring, several reviews support the use of the osmolar gap (OG) to detect mannitol accumulation; an OG around 55 mOsm/kg is a practical alert threshold that has been associated with a low rate of renal failure when respected. Therefore, OG checks alongside serum sodium and osmolality are reasonable when mannitol is administered, especially in older adults and those with baseline kidney disease. Our clinical interpretation section recommends stopping or delaying further boluses when OG approaches this range or osmolality approaches the safe threshold, with reassessment of the indication before additional therapy is delivered (41).

#### 4. Hemodynamic Effects (MAP, CPP, HR)

Mannitol causes a biphasic volume effect: an initial intravascular expansion as solute draws water into the vascular space, followed by osmotic diuresis that can reduce circulating volume hours later. Rapid infusions can acutely decrease systemic vascular resistance and cause transient hypotension, particularly at high dose rates (e.g., 1 g/kg over 10 min). These specific dosing regimens can potentially lower cerebral perfusion pressure (CPP) in patients with marginal autoregulation (41, 45, 46). Historical physiologic studies showed that rapid mannitol boluses may transiently increase cerebral blood volume, with intracranial pressure (ICP) reduction occurring only after the dehydrating phase predominates, often accompanied by a short-term decrease in mean arterial pressure (MAP) and a compensatory heart rate increase (41, 45). In contrast, hypertonic saline (HS) tends to maintain or augment MAP and CPP through more sustained intravascular expansion and sodium/osmotic effects, helping explain why HS often reduces the combined burden of elevated ICP and concomitant hypoperfusion more effectively than mannitol in severe TBI cohorts(43). Clinically, these hemodynamic dynamics support moderate mannitol infusion rates, close CPP monitoring when autoregulation is impaired, and a lower threshold to favor HS in hypotensive, vasopressor-dependent, or preload-sensitive patients.

Although individual studies have reported hemodynamic instability, such as pulmonary edema, transient hypotension, or arrhythmia, the meta-analytic summary (RR = 1.77 [0.00–6.3 × 10⁵], p = 0.67) demonstrated no statistically significant increase in cardiac adverse events associated with mannitol administration. Considerable clinical heterogeneity persisted: while Bilotta et al. (2012) observed more frequent arrhythmias and pulmonary desaturation episodes, Su et al. (2021) reported slightly fewer complications among mannitol users. Mechanistically, mannitol’s osmotic diuretic effect may reduce circulating volume and predispose vulnerable patients to transient hypotension or reflex tachycardia, whereas HS expands plasma volume and mitigates endothelial swelling factors that may underlie its comparatively cardioprotective profile in certain trials. The moderate heterogeneity (I² = 42.5%) likely reflects variation in populations and disease settings (TBI vs hepatic encephalopathy vs neurosurgical cohorts).

#### 5. Electrolyte and Osmotic Changes

The electrolyte and osmotic consequences of mannitol are central to both efficacy and risk. Early hemodilution can produce transient hyponatremia, whereas the diuresis often causes hypernatremia and hyperchloremia; potassium typically trends down with ongoing diuresis but dangerous hyperkalemia can paradoxically occur perioperatively after mannitol boluses, likely via intracellular, extracellular redistribution and effects on renal handling in susceptible patients (47). also, serum osmolality and the osmolar gap (OG) are critical considerable laboratory values. Although a historical osmolality ceiling of ≤320 mOsm/kg is frequently cited, guidelines now emphasize prioritizing OG because it correlates better with mannitol accumulation; multiple expert syntheses suggest using caution if OG rises to 20–55 mOsm/kg and avoiding further dosing when OG ≥55 mOsm/kg, recognizing that direct mannitol levels are rarely available (17, 41). These thresholds aim to reduce the risks of AKI and systemic toxicity that increase with sustained hyperosmolality. For day-to-day practice, scheduled Na⁺/K⁺/Cl⁻/Mg²⁺, osmolality, and OG checks tied to dosing frequency (e.g., pre-dose sampling when stacking boluses) and early response to derangements (e.g., proactive K⁺ and Mg²⁺ repletion) are pragmatic steps that mitigate downstream cardiac and renal events.

#### 6. Cardiac and ECG Findings

Mannitol is not uniquely “arrhythmogenic” in the absence of hyperosmolality and electrolyte disturbances. the previous literatures show adverse rhythm and ischemic events based on rate of osmolality rises, and also to ion disturbances due to dosing. Prospective work in TBI demonstrates a graded relationship between plasma osmolality and QTc prolongation, with increased risks of QTc >500 ms, ST–T abnormalities, and atrial fibrillation as osmolality climbs, sometimes at levels 310–315 mOsm/kg, lower than the classic 320 mOsm/kg ceiling (32, 40). These data confirm for conservative osmolality targets during intensive osmotherapy and support ECG/QTc surveillance when escalating mannitol exposure, especially in older adults and those with conduction disease or on QT-prolonging drugs.

Although uncommon, hyperkalemia after mannitol is a well-documented perioperative hazard that can present abruptly with peaked T waves, QRS widening, ventricular tachycardia/fibrillation, PEA, or asystole minutes to tens of minutes after a bolus. Literature reviews of such cases highlight large or rapid doses, impaired renal function, and concurrent K⁺-raising agents as contributors. Recognition is crucial because treatment (IV calcium, insulin glucose, β-agonists, bicarbonate if acidemic, ± dialysis) is time-critical. Protocols should mandate immediate post-dose electrolyte checks when dose rates are high or renal function is borderline, with preemptive potassium and magnesium optimization (e.g., K⁺ ≥4.0 mmol/L, Mg²⁺ ≥2.0 mg/dL or institutional equivalents) (47).

#### 7. Mortality, ICU Stay, and Clinical Course

Across severe TBI and mixed neurocritical cohorts, pooled analyses generally show no consistent mortality difference between mannitol and hypertonic saline (HS), despite physiologic advantages favoring HS in reducing ICP-CPP burden and, in some contexts, decreasing the need for re-intervention (35, 43, 48). intraoperative series and recent meta-analyses in craniotomy settings have associated HS with better brain relaxation and fewer “rescue” doses, which can translate into smoother intraoperative courses and, in some institutions, shorter vasopressor duration or reduced diuresis; however, these operational benefits have not reliably extended to length of ICU stay or long-term outcomes across heterogeneous populations (33, 44, 48, 49). The pooled mortality estimates in our analysis (RR =3.6 [0.83–15.6]) shows a non-significant numerical excess of deaths among mannitol-treated patients, predominantly influenced by a single small Iranian TBI trial (Hasanpour Mir 2012). That study’s unusually high mortality (79% vs 18%) likely reflects uneven baseline severity rather than a direct deleterious pharmacologic action of mannitol itself. In contrast, the larger matched-cohort and HE trial datasets (Su 2021, Hashemian 2004) demonstrated overlapping survival curves (log-rank p > 0.5). The pooled zero heterogeneity (I² = 0%) and broad 95% confidence intervals highlight the statistical uncertainty of the mortality findings and the overarching limitation of small-sample evidence (32, 40). Clinically, these results indicate no conclusive difference in survival between the two osmotic strategies. The slightly higher point estimate of risk with mannitol may reflect hemodynamic vulnerability secondary to osmotic diuresis and intravascular volume contraction, particularly in patients without aggressive volume replacement. Importantly, explicit cardiac event reporting remains sparse across most osmotherapy trials; where ECG or QTc endpoints were prospectively collected, risk appeared to track exposure factors (total osmolality, osmotic gap, infusion rate) more closely than agent identity per se, reinforcing a dose-discipline approach rather than categorical avoidance of either drug.

overally, the clinical interpretation is straightforward. When the surgical priority is brain relaxation for microsurgical exposure, HS is a pragmatic first choice, acknowledging the need to plan sodium monitoring and postoperative targets. In ICU setting, ICP crises, either agent is acceptable; when a longer effect per dose and fewer rebounds are desirable to reduce re-bolusing and nursing workload, HS is often advantageous, whereas mannitol remains entirely appropriate when sodium loading is problematic, as long as osmolality and renal indices are tracked assiduously and prophylactic use is avoided. In cardiac-vulnerable or older adults, the hazard to prioritize is over-shooting serum osmolality rather than a categorical toxicity of mannitol or HS; it is reasonable to limit osmolality to the low 300s, maintain potassium at clinically protective levels with magnesium repletion as needed, avoid stacking QT-prolonging medications, and obtain ECG monitoring during dose escalation or sustained high-dose therapy. These practical recommendations extend the framework we previously proposed for monitored, target-limited hyperosmolar therapy and formalize a context-specific approach to agent choice in the OR versus the ICU (32, 34, 50).

### What this study adds

Our work adds a specific contribution, reporting cardiac and electrolyte issues across comparative studies, domains often neglected, we provide clinically monitoring targets that can translate to bedside protocols. We supply a practical scaffold for multidisciplinary teams to decide which agent best fits a given clinical goal and risk profile. These additions build on our earlier paper, which argued for pairing ICP control with ECG/QTc surveillance and for codifying setting-specific selection rules; the present analysis widens the empirical base, extends the safety lens, and operationalizes the message with comparative tabulation. (Table 5)

**Table 5.**
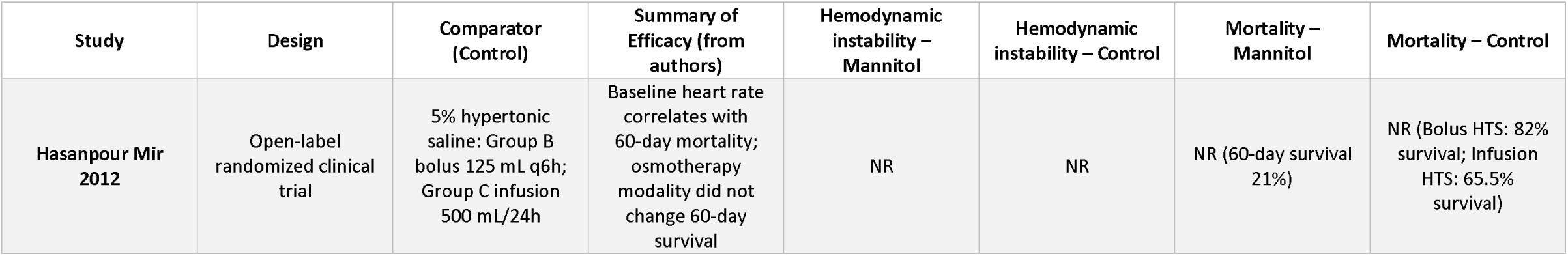
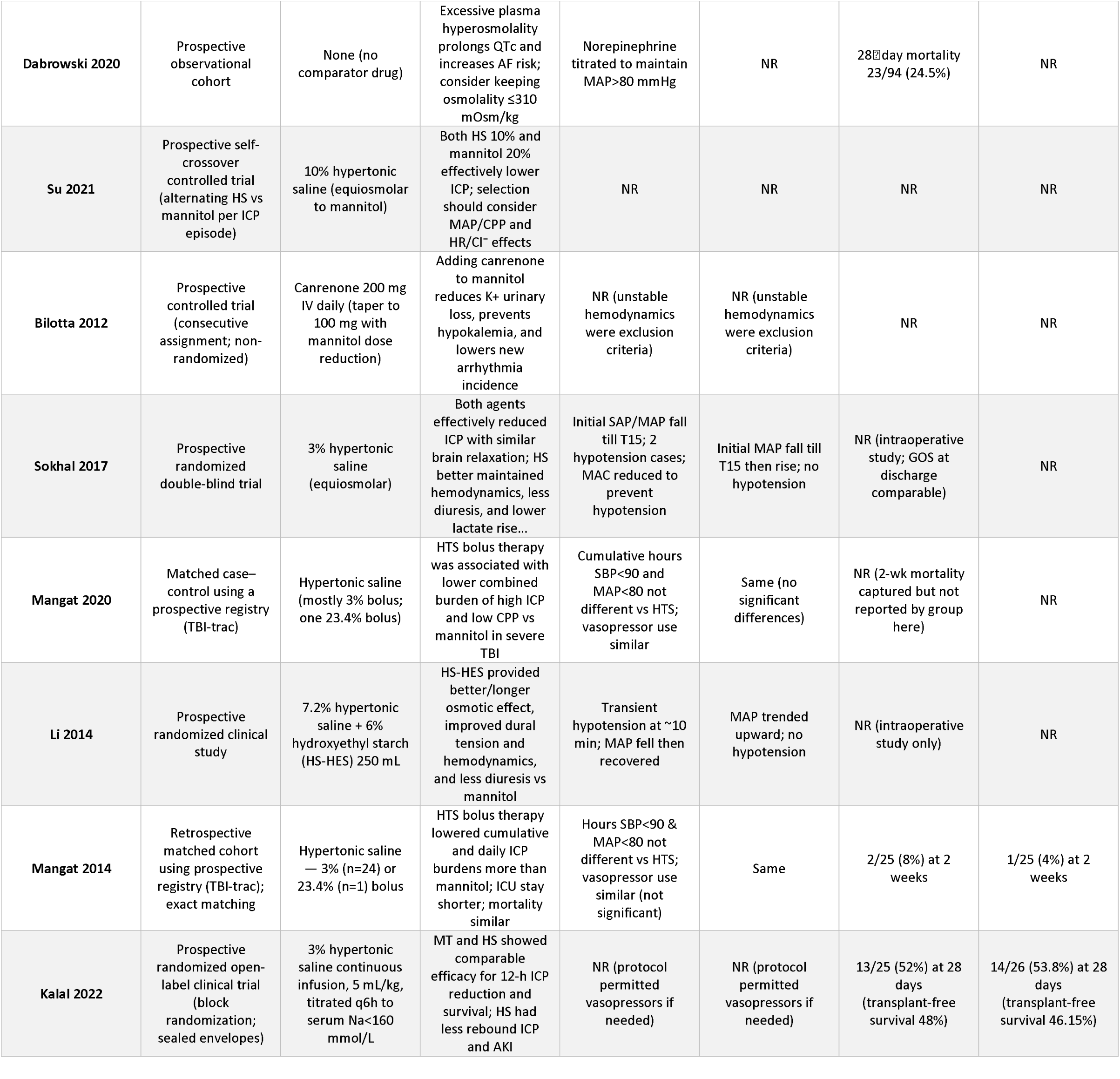
Comparative efficacy & hemodynamics (HS vs mannitol)

### Limitations

Several important limitations are present:

1. Most included studies were small, single-center trials with heterogeneous dosing strategies, varying osmolar concentrations, bolus versus infusion techniques, and inconsistent equiosmolar matching, which complicates direct comparison of mannitol and hypertonic saline. these features substantially increase the risk of performance and detection bias.
2. Critical cardiac endpoints were inconsistently reported. Few studies incorporated prospectively defined ECG monitoring, and arrhythmia detection was often sporadic, based on offline review, non-invasive surrogates, or incomplete rhythm documentation. As a result, assessments of QTc prolongation, atrial arrhythmias, and hypotension were limited by variability in measurement frequency, methodology, and adjudication.
3. Some outcomes, including mortality, long-term neurological recovery, and persistent cardiac or renal injury, were insufficiently powered or incompletely reported in most studies. Wide confidence intervals encompassing both harm and benefit reflect the statistical uncertainty of the pooled estimates. Additionally, reliance on reported adverse events without independent adjudication introduces uncertainty and a potential underestimation of cardiotoxicity or renal complications.
4. Study populations were heterogeneous, spanning neurosurgical, TBI, stroke, and liver failure contexts, with variable baseline severity and monitoring capacity, increasing the risk of residual confounding by indication.
5. The lack of individual-patient data further restricted our ability to perform stratified analyses across risk subgroups (elderly, cardiac-vulnerable patients, or those with impaired renal reserve).
6. Finally, small hemodynamic datasets raise the possibility of publication bias, particularly because studies with neutral or negative safety findings may be less likely to have been published.

### Future Directions

Future trials must incorporate integrated ICP-control metrics together with standardized cardiac and renal safety:

1. cardiac monitoring:
2. ECG at baseline, post-dose, and during dose escalation
3. Predefined QTc thresholds triggering dose modification
4. Blinded adjudication of arrhythmias
5. Standardized biochemical laboratory tests:
6. periodic checks of electrolytes (Na⁺, K⁺, Mg²⁺, Cl⁻)
7. Serum osmolality monitoring
8. Osmolar-gap thresholds
9. Patient-based outcomes:
10. ICU and hospital length of stay
11. Long-term functional outcomes
12. Absolute risk differences in cardiac events and acute kidney injury
13. Harmonization of key definitions:
14. Consistent scales for brain relaxation
15. Standardized definitions of rebound ICP
16. Adoption of time-integrated ICP/CPP burden as a primary or secondary outcome. Conclusion:

Mannitol provides effective, rapid ICP reduction with minimal and transient hemodynamic impact, showing no significant increase in cardiac adverse events or mortality compared with hypertonic saline across nine studies involving 428 patients. Observed cardiac adverse events, including QTc prolongation and electrolyte disturbances, were more closely related to elevated serum osmolality than to the hyperosmotic agent itself. Although based on methodological limitations, current evidence supports the cardiovascular safety of mannitol when administered with appropriate monitoring. A precautionary approach emphasizing serum osmolality <320 mOsm/kg, osmolar-gap variable, and structured ECG and electrolyte monitoring remains considerable and necessary. Robust multicenter RCTs with standardized cardiac endpoints are needed to refine risk estimates and guide optimal osmotherapy selection.

## Data Availability

All data produced in the present study are available upon reasonable request to the authors

## Acknowledgments

The authors have no acknowledgments to declare.

## Appendix and Supplementary Material

1. Supplementary 1. Search Strategy
2. Supplementary 2. Title abstract Exclusion sheet
3. Supplementary 3. Full text screening Exclusion sheet
4. Supplementary 4. Data extraction Sheets
5. Supplementary 5. Risk of bias Tables

